# The Food, Activity, Screens, and Teens (FAST) Study; Design and protocol

**DOI:** 10.1101/2024.10.08.24314237

**Authors:** John R. Sirard, Lindiwe Sibeko, Mark C. Pachucki, James A. Kitts

## Abstract

The Food, Activity, Screens, and Teens (FAST) Study was a school-based prospective cohort study, aiming to identify mechanisms of peer influence on weight-related behaviors (WRBs) among early adolescents. In 2017-18, FAST investigators conducted focus group interviews and field observations of sixth grade students at four ethnoracially diverse urban middle schools, then administered pilot surveys in two of the schools. In Fall 2018, investigators recruited a cohort of sixth graders in the same four schools for a three-year panel study, with four waves per year. Each wave measured height and weight, demographic characteristics, WRBs (physical activity, screen time, and dietary patterns), classes and activities, and social networks among peers. Peer network measures included friendship, social sentiments (liking/disliking), online and face to face interaction, kinship, cohabiting, and shared WRBs. The pandemic school closure in March 2020 interrupted fieldwork after wave 6, and the next five waves employed online and mail surveys while schools operated remotely. In Spring 2022, after schools reopened, investigators followed a subset of students into high school to collect a twelfth wave of data.

## INTRODUCTION

Overweight/obesity is often clustered among friends and there is growing evidence for social influences on adolescent weight status [1–10]; If there is an apparent social contagion underlying the epidemic of overweight/obesity, it presumably operates through antecedent weight related behaviors (WRBs), such as poor dietary choices [11], low levels of physical activity [12] and high levels of screen media use [13, 14]. These behaviors are initially shaped by parent and sibling modeling [15–17], and may then be modified by the increasing influence of peers[18] in the broader context of health lifestyles [19]. Prior findings of social causation for WRBs have been inconsistent given vexing inferential challenges, however [18, 20–22]. For example, clustering of friends with similar WRBs could be due to peer *influence* on behaviors, but may also be due to *selection* of friends with similar WRBs, or exposure of friends to similar external *contexts* affecting WRBs, and may be partly a byproduct of triad *closure*, the tendency for friends of friends to become friends [23]. Application of experimental methods to identify these interdependent forces has been challenging due to an inability to randomly assign friends and friends’ WRBs within empirical contexts and on time scales where these processes naturally operate.

The Food, Activity, Screens, and Teens (FAST) Study was a prospective cohort study that collected longitudinal data on demographics, weight-related behaviors (physical activity, screen time, dietary patterns), height and weight, classes and extracurricular activities, as well as social relationships and social interaction among adolescents over an extended period. We aimed to deliver insights into the interplay between health behaviors, weight status, and peer relationships over time in a racially and ethnically diverse sample of youth in four urban middle schools.

Earlier work on youth relationships and health behavior in schools largely examined friendships [18, 20, 24–28], but recent work [29] has criticized this concept as ambiguous and argued that it conflates the role relation of friendship with alternative concepts such as interpersonal sentiments (e.g., liking) and social interaction (sharing social time and activities, including WRBs). The FAST Study goes beyond earlier work to distinctly measure role relations (e.g., friendship and kinship), interpersonal sentiments (from disliking to liking), general measures of interaction (e.g., spending time together, online and face-to-face), and more specific forms of interaction that involve shared WRBs (e.g., shared eating, physical activity, and screen time). This distinction will allow unique insight into the mechanisms of peer effects through modeling the underlying selection and influence processes on adolescent overweight/obesity and antecedent WRBs [21, 28, 30].

## METHODS

### Design

The FAST Study enrolled longitudinal, observational cohorts of youth in four middle schools, partnering with school and district leadership. Data collections were planned four times each school year (September, December, March, June) for three years (6^th^ grade [September 2018] to 8^th^ grade [June 2021]). The large number of waves and high frequency of data collection (approximately every three months) offered additional information for identifying the interplay between peer effects and several contextual and psychosocial factors on WRBs over time. Importantly, the measurement timing coincided with largely exogenous changes in the environment (return from summer vacation, class schedule changes, athletic seasons) that influence interaction opportunities among school-aged peers. These shifts in opportunities for interaction and activity may indirectly affect interpersonal relationships and WRBs.

At the midpoint of the study, the COVID-19 pandemic resulted in closure of schools, clubs, gyms, and restaurants, and led to widespread social distancing. Although the closures and distancing in 2020 proved challenging for our data collection, they also offered a substantial exogenous intervention to social interaction networks, allowing us to examine pandemic effects on friendships, sentiments, shared activities, and WRBs.

Due to the COVID-19 pandemic closure, data collection was temporarily halted in March 2020, days before scheduled administration of wave 7. This resulted in a missed survey for March of 2020, which was made up with a remote survey (both a web survey and paper survey sent by mail) administered in August, 2020. Although late, wave 7 used a retrospective period of self-reported interaction and health behavior for January 2 until the school closure, March 13, 2020, closely matching the period originally intended for wave 7. Due to school closures, wave 8 was another remote survey (again both a web survey and a mailed paper survey) in October 2020, with a retrospective period of self-reported interaction and health behavior for the entire period when schools were closed from March 13, 2020, until the start of school in September, 2020. Wave 8 thus matches the period originally intended for waves 8 (Spring) and 9 (Summer). The next three waves (waves 9, 10, and 11) were implemented on schedule for Fall, Winter, and Spring of the students’ 8^th^ grade year. These surveys were administered online and by mail during the period of remote schooling. As schools were beginning to reopen during wave 11, surveys were also administered in-person for some students at one school. A twelfth wave was implemented in-person a year later (June of 9^th^ grade) for participants who could be reached in high school (Figure 1).

**Figure 1:**
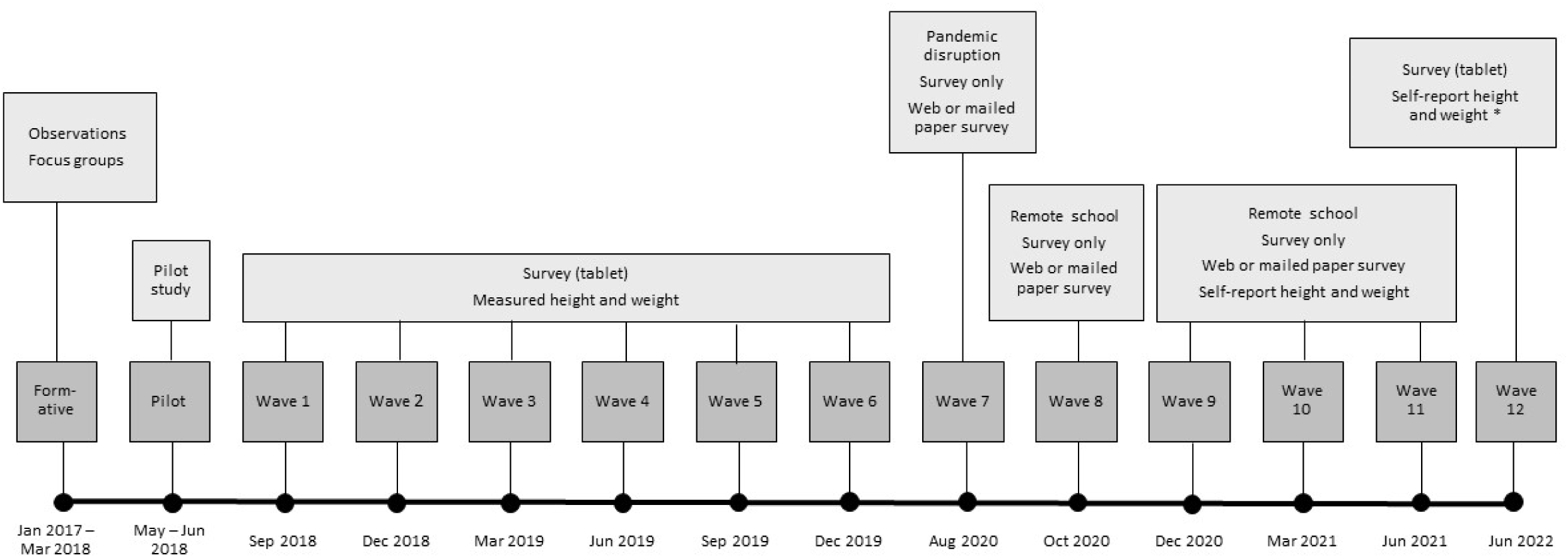
Fast Study timeline. *Measured height and weight was available forn=19 studentsat one school

### Schools and Student Participants

To protect the privacy of participants and schools, specific locations are not provided and school names have been changed. All study procedures were approved by the university Institutional Review Board and by the partnering school district. The school district, located in the Northeast region of the United States, is predominantly low income (all students received free breakfast and most received free lunch) and non-White (Hispanic/Latino and Black/African American). Four public middle schools (6^th^ to 8^th^ grade) participated in this study. The demographics at the four partnering schools are provided in Table 1. The smallest cohort was at an arts-focused magnet school (ARTS, N = 55 at wave 1) and had a majority female (72%) population. The largest cohort was at a magnet school with a focus on science and technology (TECH, N = 99 at wave 1, 43% female). The third cohort was at a college-preparation focused magnet school (PREP, N = 92 at wave 1, 54% female). The fourth cohort was at a traditional neighborhood school serving a specific geographic area of the city (EAST, N = 71, 50% female).

**Table 1:** Baseline characteristics for 6^th^ grade students at participating schools and averaged 517 across schools (anonymized)

| **School** | **(N)**<br>**6<sup>th</sup> grade** | **%**<br>**Low Income** | **% non-**<br>**White** | **%**<br>**Black/AA** | **%<sup>518</sup>**<br>**Hispanic** |
| --- | --- | --- | --- | --- | --- |
| PREP (Magnet) | 92 | 55 | 85 | 22 | 61<sup>519</sup> |
| EAST (Neighborhood) | 71 | 96 | 94 | 10 | 88 |
| TECH (Magnet) | 99 | 71 | 87 | 16 | 67<sup>520</sup> |
| ARTS (Magnet) | 55 | 79 | 87 | 20 | 66 |
| All Participating Schools | 317 | 75 | 88 | 17 | 70.5 |

Data from: State Department of Education
Black/AA = Black/African American
Low income: Calculated by the state Department of Education. Based on a student's participation<sup>2</sup> in one or more of the following state-administered programs: the Supplemental Nutrition Assistance Program (SNAP); the Transitional Assistance for Families with Dependent Children (TAFDC); the Department of Children and Families' (DCF) foster care program; and Medicaid.

### Formative Work and Pilot Study

Formative work lasted from January 2017 to March 2018. During this period, we interviewed school administrators to learn about the school’s policies, practices, and infrastructure. Direct observation of students provided general information on culture and norms at each school. Focus groups were used primarily to develop, test, and revise survey questions. At least two focus groups involving, on average, four students per group were conducted at each school, in both English and Spanish, segregated by gender. Focus group participants went through an active written informed parental/guardian permission process and students provided their written assent at the beginning of the group sessions (February to March 2018).

From May to June 2018, the Pilot Study was conducted at two of the four participating schools; the traditional neighborhood school (EAST) and one of the larger magnet schools (PREP). We measured height and weight, and administered our draft survey, encouraging students to ask clarifying questions. We revised the survey based on participant questions and detailed field notes so that the survey plus height and weight measurements could be completed within a 50-minute classroom period. The students for the formative work and pilot study were in the 6^th^ grade in 2017-2018, and thus were different from the students enrolled in the prospective cohort study.

The formative work (i.e., observations, focus groups, and interviews) was crucial in helping us develop and revise survey questions, but also later aids in interpreting responses and identifying anomalies.

### Consent and Assent Processes

In September 2018 we invited all 6^th^ graders at the four partnering schools to participate in the study. To achieve the high response rate needed for meaningful network analysis [31], participants were recruited using a written passive informed consent process, where parents/guardians were notified by letter and all students in each cohort were allowed to participate unless their parent/guardian replied with a refusal. The school district agreed to the passive consent process with the understanding that survey questions would not involve potentially sensitive topics (e.g., child abuse, alcohol or drug use, sexual activity, disordered eating, bullying and teasing, body image, mental health).

In addition to the consent process, students provided their written assent to participate in the study at the beginning of each wave of data collection. Each data collection session began with a brief description of the study and review of the assent form prior to beginning the survey or having their height or weight measured. Students could assent and complete the survey but refuse to have their height and weight measured. If new students joined the school during the duration of the study (September 2018 to June 2021), but after this initial recruitment, the same procedures for parental/guardian permission and student participant assent were followed.

Students whose parents opted them out of the study (n = 15, 4.7%) and those that refused assent in a given wave were instructed to read or work on material provided by the teacher. Sample sizes at each data collection wave are presented in Table 2. Lower participation rates in waves 8 through 11 are due to remote survey administration during the pandemic, and in wave 12 due to inability to access students at some high schools (9^th^ grade year). All recruitment, consent, and assent procedures were approved by the university’s Institutional Review Board and the participating school district.

**Table 2:** Participation rates by school and wave (% participation, number of participants / total enrollment in the grade)

| Wave | ARTS | EAST | PREP | TECH |
| --- | --- | --- | --- | --- |
| **Pilot** |  | 90%, 69/77 | 84%, 82/98 |  |
| **01** | 84%, 46/55 | 88%, 64/73 | 90%, 93/103 | 95%, 88/93 |
| **02** | 87%, 48/55 | 90%, 65/72 | 94%, 98/104 | 96%, 94/98 |
| **03** | 84%, 46/55 | 84%, 63/75 | 93%, 93/100 | 90%, 88/98 |
| **04** | 82%, 45/55 | *67%, 51/76 | 91%, 91/100 | 84%, 83/99 |
| **05** | 83%, 44/53 | 91%, 74/81 | 91%, 91/100 | 88%, 85/97 |
| **06** | 85%, 44/52 | 80%, 67/84 | 90%, 88/98 | 88%, 88/100 |
| **07\*\*** | 21%, 11/52 | 16%, 14/86 | 27%, 27/99 | 24%, 23/96 |
| **08** | 81%, 44/54 | 67%, 65/97 | 83%, 81/98 | 65%, 64/98 |
| **09** | 71%, 37/52 | 79%, 73/92 | 78%, 75/96 | 70%, 64/92 |
| **10** | 71%, 37/52 | 73%, 68/93 | 80%, 77/96 | 51%, 45/88 |
| **11** | 56%, 29/52 | 64%, 60/94 | 73%, 69/95 | 45%, 40/88 |
| **12** | 57%, 31/54 | 34%, 33/96 | 25%, 24/96 | 40%, 37/92 |

**Notes:**
\* Low participation rate largely due to a single class, where the teacher allowed students to play a video game (instead of reading) as an alternative to participating in the study.
\*\* Low participation rates due to Covid school closure and mailed survey distribution during summer 2020

### Measures

#### Sociometric Constructs

Previous work on adolescent social networks has focused primarily on *friendship* networks, interpreting friendship networks as synonymous with networks of social interaction or sentiments [29]. Our focus groups revealed that adolescents in this population defined friendship primarily in terms of normative obligations, and derived a set of 11 descriptors that our subjects use to define the friendship concept. Throughout the study, the electronic survey allowed respondents to rank those 11 descriptors (presented in initially random order) in importance for defining friendship. Analysis of pilot study data [32] confirmed that friendship is primarily a normative concept, and found that girls and boys use the term differently. Our pilot study also showed that direct measures of social *interaction* (spending time together in person or online), social *sentiments* (liking or disliking), and *access* to one another (shared classes, neighborhoods, clubs) produce networks that are quite distinct from networks of *role relations* like friendship or kinship. Even questions about different forms of interaction (e.g., face to face vs. online interaction, lunch cafeteria partners, physical activity partners) elicited clearly distinct networks. Therefore, the FAST Study network measures captured these four distinct network concepts [33] rather than merely measuring friendships.

We operationally defined *access* as the participants’ class schedules and residential proximity to each other, using data provided by the school district, and measured cohabiting and shared club or team membership using our survey. The remaining network measures were derived from our survey. We measured *role relations* as self-reported friendship or kinship around the time of the survey. We measured *sentiments* around the time of the survey on a 5-point scale from Strongly Dislike to Strongly Like, with neutral as the default choice. Our *General Interaction* questions assessed whether the participants spent time together (face to face or online) at least once per *week* during the recall period of each wave (normally three months). Our *Health Behavior Interaction* questions assessed interaction specifically related to the target health behaviors – physical activity, screen time, eating – which are explained in the survey. These behaviors are a narrower subset of total interaction, so we used a more liberal interaction threshold (at least once per *month* during the recall period of each wave). We also asked respondents to report with whom they regularly sit in the cafeteria. The key wording for the 10 network questions is summarized in Table 3 and full wording for all questions is provided in Appendix A.

**Table 3:**
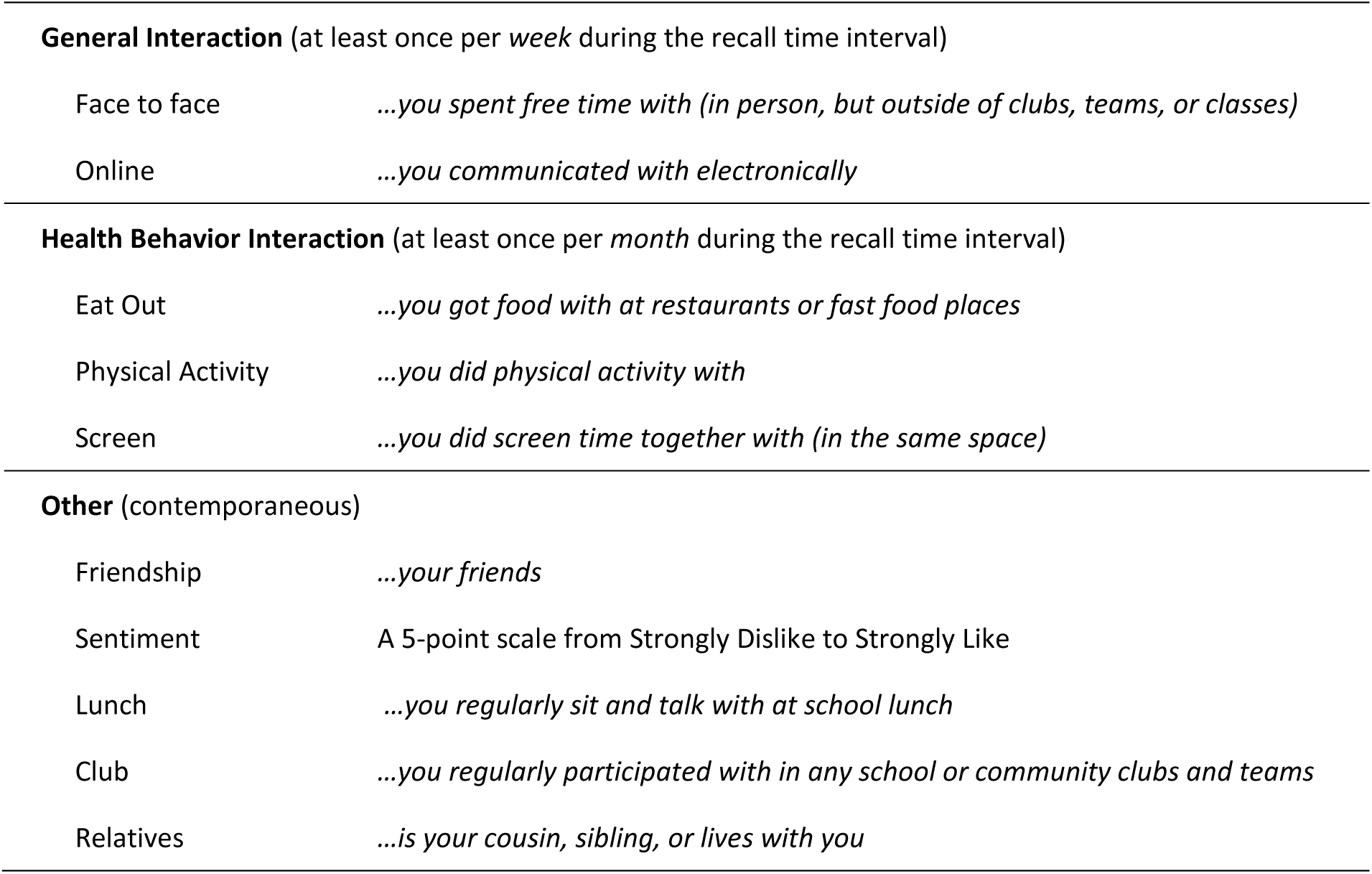
Key wording for the 10 Sociometric Questions.

|  |  |
| --- | --- |
| **General Interaction** (at least once per week during the recall time interval) |  |
| Face to face | *...you spent free time with (in person, but outside of clubs, teams, or classes)* |
| Online | *...you communicated with electronically* |
| **Health Behavior Interaction** (at least once per month during the recall time interval) |  |
| Eat Out | *...you got food with at restaurants or fast food places* |
| Physical Activity | *...you did physical activity with* |
| Screen | *...you did screen time together with (in the same space)* |
| **Other** (contemporaneous) |  |
| Friendship | *...your friends* |
| Sentiment | A 5-point scale from Strongly Dislike to Strongly Like |
| Lunch | *...you regularly sit and talk with at school lunch* |
| Club | *...you regularly participated with in any school or community clubs and teams* |
| Relatives | *...is your cousin, sibling, or lives with you* |

For each network question, the content of the question was followed by a roster containing the names of all same-grade students. Respondents could select, “There is no one on this list that I want to select” to allow us to differentiate a non-response from a lack of ties. Importantly, there was no limit on the number of students that could be nominated for any of the network questions. For the teams and clubs question, a series of follow-up questions aimed to identify the names of teams or clubs that the respondent participated in jointly with each nominated peer. For the “relatives” question, a series of follow up questions aimed to clarify the specific relationship of the nominated peer to the respondent (i.e., sibling, cousin, or housemate).

Interaction questions were asked before the friendship question to avoid priming participants to recall just those they consider friends, aiming to capture a broader range of interaction partners.

Similarly, general interaction questions were asked before health behavior interactions to avoid priming just those who share health behaviors. Health behavior interaction questions were presented in random order for each subject in the electronic survey. Preliminary analyses [34] revealed no evidence of bias due to the order of names on the roster. Randomization of question order for health behavior interaction questions allowed us to examine question order effects, and preliminary analyses indicate no evidence of question order effects. To assess the extent of student friendship networks outside of cohort peers, the survey also asked about the number of friends in other grades or at other schools.

#### Weight-Related Behaviors

Questions that assessed WRBs of interest (physical activity, screen time, dietary patterns) and sleep patterns were modified from existing validated instruments. Most behavior questions were retrospective to the period since the previous survey (typically 3-4 months). See Appendix A for question wording.

*Physical Activity* questions were modified from the 3-item Godin-Shepard Physical Activity Survey [35] that has been previously used in middle and high school students [18, 36, 37]. Prior to any physical activity question (including the social interaction question related to physical activity), a definition was provided to ensure that respondents were considering all physical activity, not just organized sports. Questions asked about vigorous, moderate, and light intensity physical activity with relevant examples for each, derived from the focus groups and pilot testing.

*Screen time* was assessed with five items that have previously been used in adolescent populations and, consistent with previous studies, the same questions were asked separately for weekdays and weekends [38, 39]. Participants recalled times when they engaged in these screen behaviors while lying down or sitting, to capture sedentary screen time. The questions reflected new technologies and focused on the types of screen behavior (e.g., TV/Streaming, video games, social media, voice/video communication, web browsing) regardless of the platform used (phone, tablet, or computer). Two additional questions captured whether respondents played active video games, “where you’re standing up, dancing, jumping.”

*Dietary patterns* were assessed with a series of questions that focused on frequency, during a regular week, of healthy and unhealthy dietary patterns that have been associated with healthy weight or excess weight gain in youth [38, 40, 41]. Questions included frequency of eating takeout/delivery meals and snacking during a regular week. Frequency of drinking eight types of beverages including: regular soda, diet soda, regular sports drinks, sugar-free sports drinks, other sugar-sweetened beverages, other sugar-free beverages, 100% fruit juice, and water. Breakfast consumption during a regular week was assessed with one question, and two questions assessed frequency of fruit and vegetable consumption during the past week. Lastly, one binary (yes/no) question determined if the participant had “gone on a diet to lose weight” during the wave period.

*Sleep* patterns were assessed in waves 10-12, in response to the Covid pandemic, using questions from the Sleep Habits Survey for Adolescents [42]. Questions were asked separately for weekdays and weekends, to determine what time participants usually went to bed in the evening, what time they usually got out of bed in the morning, and how many nights they had trouble falling asleep.

#### Psychosocial Variables

##### Social Support (Peers

At each wave, the survey assessed the level of social support participants received from their friends for physical activity (three questions) [15, 36]. At the first wave that each participant responded, the survey also asked about the social support participants received from their parents for physical activity (two questions), eating a healthy diet (four questions), and limiting their screen time (two questions), plus social support for physical activity received from older siblings (two questions) [15, 36, 43].

##### Subjective social status

A ladder-type question assessed participants’ perceived popularity in the school (modified from Goodman et al. [44]), whereas the social network data (e.g., friend and sentiment measures) provided independent measures of participants’ popularity within the grade cohort.

#### Demographics

The survey asked participants to report their sex (male or female), birth date and race/ethnicity (*American Indian or Alaska Native, Asian, Black or African American, Hawaiian or Other Pacific Islander, Hispanic or Latino,* or *White*). Multiple race-ethnic categories could be selected. A follow-up question reminded each participant of the full set of their race/ethnicity responses and then invited them to elaborate on or explain their responses in an open text field. Starting in wave 6, participants could choose “Other” and elaborate in an open text field, and an additional question asked about finer-grained identities derived from open text fields in earlier waves (*Brazilian, Dominican, Puerto Rican, Salvadorian, Venezuelan*). From wave 7 onward this question was expanded to (*Bahamian, Brazilian, Cuban, Dominican, Guatemalan, Jamaican, Mexican, Panamanian, Peruvian, Puerto Rican, Salvadorian, Venezuelan*) and the option “*Hispanic or Latino*” was broken into 2 choices: “*Hispanic*” and “*Latino*.” From wave 10 onward, participants were asked in a separate question if they identified as “*Multiracial or Mixed Race*.” Starting in wave 4, participants indicated all languages they speak and the language they speak most often at home.

#### Onset of Puberty

To assess onset of puberty, girls were asked about age at menarche and boys were asked about age at voice deepening and facial hair growth [45, 46]. The puberty questions were asked at each wave until a student responded in the affirmative.

#### COVID-19 Questions

For the four data collection waves during the 2020-2021 academic year, the survey included additional questions related to participant experiences with COVID-19 (whether they had been diagnosed with or tested positive for COVID-19, had a known exposure to COVID-19, or wished to be vaccinated). An open-ended question asked about how the pandemic was affecting their social lives and WRBs. During these waves, survey questions also asked about participation in online PE classes and working on PE assignments outside of school hours. Given mandates for social distancing during the early pandemic, two questions asked about the *amount of time* spent interacting with others outside of school hours; separate questions were asked for interacting with other cohort members and interacting with students who go to other schools (or are in other grades at their school). Separate questions for face to face and online interactions were also included. During school reopening (wave 10), one question asked if the *number of people* they interacted with face to face had increased, decreased, or stayed the same.

#### Household Characteristics

The survey included questions to capture basic household characteristics, such as household size, number of older and younger siblings, and which relatives and non-relatives lived in the household. To assess youth perceptions of family socioeconomic status, the survey also included the Family Affluence Scale [47, 48] with minor modifications for the contemporary era, asking subjects to report the following about their household: number of bathrooms at home, having one’s own bedroom, having a clothes washing machine at home, number of computers, vacation travel, and number of vehicles owned. The Wave 11 and 12 surveys also asked for education and employment status of parents in the subject’s primary home (father or stepfather, mother or stepmother). Questions about parental education and employment were not asked earlier due to results from our pilot study indicating that the 6^th^ grade students struggled to respond, with just over half the responses being “I don’t know”. Further, heterogeneous family structures complicated measurement of respondents’ family social support, socioeconomic status, parental education and employment, siblings, and who the respondent lived with. We sought to develop questions that would be generally applicable to respondents from diverse households without being overly complicated.

#### Height, Weight

For waves 1 to 6 (before the pandemic school closure), each student’s height and weight were measured by trained research staff to the nearest 0.1 cm and 0.1 kg, respectively, using a portable stadiometer (Weigh and Measure, LLC, Olney, MD) and digital scale (Seca 876, Hanover, MD). Students were measured in a semi-private location away from the survey administration and without shoes, coats, jackets, and heavy sweaters, or sweatshirts. Participants could see the scale display but it was in kilograms, not pounds, and if students inquired about their height or weight they were encouraged to look up the conversion factors online. During waves 9-12 two questions were added to the survey to allow participants to self-report height (in feet and inches) and weight (in pounds). In-person measurement was permitted at one middle school in wave 11 and at two high schools in wave 12. For that subset of the population, both self-report and measured height and weight are available.

### Data Collection Protocol

The FAST Study survey mode of administration was adapted to the pandemic school closure (March 2020) and subsequent full reopening of schools (September 2021). The survey was administered in school via digital touchscreen tablets (waves 1-6, 12), remotely using web surveys (waves 7-12), or mailed paper surveys with self-addressed stamped envelopes (SASE) (waves 7-11). Both tablet and web surveys employed Qualtrics software (Qualtrics, Provo, UT). Most data were collected during school hours. Data were collected at each school on a single day (with make-up days for absent students). The data collection days for the four schools were scheduled around the same time, at least within a 2-week period for each wave.

Trained research staff prepared tablets for survey administration. Desks were spread apart as much as possible to increase privacy and discourage talking during the survey. Tablets had privacy screens to prevent participants from seeing each other’s responses. Research staff provided brief scripted instructions followed by the assent. Participants were allowed to work at their own pace and raise their hand if they had a question. Participants could skip questions but, for most questions, were prompted with a pop-up screen with options to complete the question or continue without answering. After the survey and height and weight measurements, participants received a $10.00 gift card to a national retail store for participating in each data collection session. At least one makeup session was scheduled at each school to collect data from those who were absent on the main day. To encourage continued participation during the pandemic, additional raffles were held in each school for headphones and $100.00 and $200.00 gift card prizes. The gift card amount for participation was increased to $20 for wave 12 to further incentivize participation.

For wave 7 (several months after the pandemic closed schools) participants were sent a letter with a link to a web-based version of the survey and a paper copy of the survey with a self-addressed stamped envelope; students chose how they wanted to respond. After classes began operating in virtual classrooms in September, 2020, from wave 8 to wave 11 most students responded to the survey during a synchronous online classroom with a survey link provided in the chat window of the virtual classroom. Students who were absent on the time of survey administration were sent a packet containing an invitation letter with instructions on how to access the web survey, a paper copy of the survey with a SASE for returning the paper survey if they chose this mode. If students did not respond to the survey after two weeks (waves 7-11), they were sent a one-page reminder letter. If there was still no response after another two weeks, a final reminder letter was sent with web survey instructions, paper survey and SASE. The survey was revised as necessary for the hardcopy mail survey, and questions that required the Qualtrics software interface were dropped.

### Quality Control

Respondents’ self-reported date of birth was reconciled across all waves and checked against school district administrative data, then used to compute age at each data collection wave. Respondent sex was reconciled across all waves and checked against school district administrative data. Multiple data checks and identity verifications allowed us to detect and later fix anomalies, such as when participants responded to the survey more than once, selected the wrong name for themselves in the survey, or (in one vexing case) two participants swapped tablets in mid-survey. We were able to identify some low-quality survey responses, such as when a subject selected the same choice for an entire block of questions, or claimed to be friends with everyone, when that claim was not corroborated by other participants and was very inconsistent with observations from other waves.

Duplicate survey observations (e.g., when a subject completed a survey twice, responded both to an online and paper survey, or stopped in mid-survey and then started over at a later time) were compared and reconciled. Duplicate measures often included low-quality values, where a participant skipped through questions seen a second time. Thus, for duplicate values on questions employing selection among fixed choices, we took the *first* response value unless it was missing or marked as suspect for other reasons. For questions employing multiple checkboxes (e.g. race identities, network tie nominations), we took the union of responses across duplicate observations.

Use of multiple measures and multiple temporal observations also provide information about subjects’ response patterns and facilitate identification and interpretation of response errors. For example, repeated measurement and subsequent refinement of demographic questions along with open text fields that allowed participants to explain their answers gave us opportunities to observe, diagnose, and correct reporting errors by respondents in the data cleaning process. We liberally used open text fields to provide context for checkbox responses and identify misunderstandings (e.g., Caribbean students who once mistakenly checked “Pacific Islander” or Puerto Rican students who once mistakenly checked “American Indian” when they had meant *American*.)

Similar to survey responses, anomalies in height and weight measures were reconciled using multiple waves of available data and field notes taken during the data collection sessions. Duplicate measures of height and weight were averaged.

Use of multiple measures and multiple observations also allowed for cleaning of the sociometric data. Notably, the interaction questions are symmetric by design (i.e., if subject A has lunch with subject B, then B must also have lunch with A). Measures of whole networks (rather than capturing ego networks for a sample of individuals) enables comparing reports by both partners. This allows diagnostic work [49] to adjust for errors in self-reports of interaction partners, such as egregious over- or under-reporters of network ties.

## CONCLUSION

The aim of the FAST Study is to better understand the social network processes that potentially shape WRBs in middle school students. By better understanding these processes, our long-range goal is to improve efficiency and effectiveness of WRB interventions by incorporating social network information into the design and implementation of these interventions.

## Data Availability

All data produced in the present study are available upon reasonable request to the authors.

## ACKNOWLEDGEMENTS

The authors sincerely thank the school district, school administrators, teachers, staff, and students for their cooperation and participation in this research project.

## APPENDIX

Survey Questions from Wave 11 (June 2021; end of the cohort’s 8^th^ grade year)

Note: The phrase, “ since around late March” that appears in a number of questions is specific to this wave of data collection and the wording was different for each wave to ensure students were recalling the correct timeframe.

1. Check the box next to YOUR name from the list below.
*Names are listed in alphabetical order by first name*.
*If you cannot find your name on this list, check “**I can’t find my name on the list above!**” at the bottom and then write your name in below.*

<<ROSTER>>

**□ I can’t find my name on the list above!**

If you’re sure your name isn’t on the list above, write it in here (first & last).

____________________________________

2. **Think about the time since we last gave this survey** (around late March). Pick the 8th graders that you **<u>spent free time with</u>** (in person, but outside of clubs, teams, or classes) **<u>at least once per week</u>**since around late March.

<<ROSTER>>

**□ There is no one on this list I want to pick.**

3. Pick the 8th graders that you **<u>communicated with electronically in your free time at least once per</u> <u>week</u>** since around late March. This means you chat back and forth with each other by voice, text, or video on a device (computer, tablet, phone).

<<ROSTER>>

**□ There is no one on this list I want to pick.**

4. There are probably some people in your grade that you truly like, some that you truly dislike, and many you have no real opinion about either way. Please tell us about this using the following check boxes.

**<u>(You don’t need to respond about everyone</u>**. You can leave many of them blank, and just check the boxes for the people you truly like or dislike.)

| Student | Strongly dislike | Dislike |  | Like | Strongly like |
| --- | --- | --- | --- | --- | --- |
| First & Last Name | ☐ | ☐ | ☐ | ☐ | ☐ |
| First & Last Name | ☐ | ☐ | ☐ | ☐ | ☐ |
| Etc... |  |  |  |  |  |

5. Think about the students that you **<u>like</u>** in the 8th grade at your school. For you, how important is it that they **<u>agree with you</u>** on who else you like (and dislike) in the 8th grade at your school?

□ Not at all important
□ Slightly important
□ Moderately important
□ Very important
□ Extremely important

6. (* Note, the electronic version of this question used skip logics based on participant responses. The version presented here was used for the paper version of the survey.)

**Think about any organized group such as** school or community clubs and teams that you regularly participated in since around late March. Please also include any organized group even if the meetings were online due to the shutdown.

*Check the boxes to the left of the 8th graders* that you **regularly participated with in these organized groups – school or community clubs and teams (including online) -- since around late March.** Don’t worry about the circles right now.

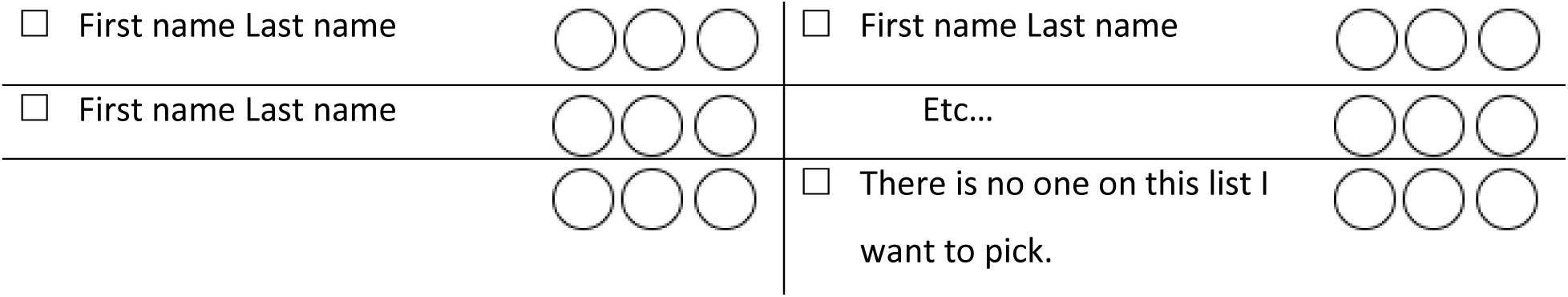

|  |  |  |  |
| --- | --- | --- | --- |
| ☐ First name Last name | ○ ○ ○ | ☐ First name Last name | ○ ○ ○ |
| ☐ First name Last name | ○ ○ ○ | Etc... | ○ ○ ○ |
|  | ○ ○ ○ | ☐ There is no one on this list I want to pick. | ○ ○ ○ |

On the lines below, write in the names of the **organized groups** – school or community clubs and teams -- that you participated in with any of the 8th graders that you checked on the previous pages.

Organized Group #1: ________________________________

Organized Group #2: ________________________________

Organized Group #3: ________________________________

Organized Group #4: ________________________________

Organized Group #5: ________________________________

For organized groups you listed above, see the **Organized Group number (#)** to the left of each organized group. **Write this number into the circle** on the previous page, **next to the name(s) of the 8^th^ graders who share that organized group with you.**

7. You may also participate in organized groups where no other 8th graders also participate. What were the <u>organized groups</u> that you **<u>participated in regularly since around late March</u>,** where <u>no other 8th graders from your school</u> participated in that organized group?

If none, write “none” in the space below.

__________________________________________________

8. Think about the times you went with other 8^th^ graders to get food from restaurants or fast food places (McDonalds, Burger King, KFC, Dunkin Donuts), Walgreens, Dollar Store, convenience stores, bodegas, or gas stations (Example: Valeros, Pride).

Check the boxes next to the 8th graders that you **got food with** at these places **at least once per month since around late March.**

<<ROSTER>>

**□ There is no one on this list I want to pick.**

9. **PHYSICAL ACTIVITY** is any game, sport, or exercise that makes you move faster and breathe harder; makes your heart beat faster; and makes you sweat a little or a lot.

Check the boxes next to the 8th graders that you **did physical activity with at least once per month since around late March.**

<<ROSTER>>

**□ There is no one on this list I want to pick.**

10. **SCREEN TIME** is watching TV/DVDs/streaming, playing video games (on a console, computer, tablet/phone), using social media or other computer software, including apps for voice, text, and video chatting.

Check the boxes next to the 8th graders that you **did screen time together with** (in the same space) **at least once per month since around late March.**

<<ROSTER>>

**□ There is no one on this list I want to pick.**

11. Think about where you sit during school lunch. Pick the 8th graders that you **<u>regularly sit and</u> <u>talk with during school lunch.</u>**

<<ROSTER>>

**□ There is no one on this list I want to pick.**

11a. Feel free to tell us anything to explain your choice about who you sit with at school lunch, in the space below.

_______________________________________________________________

12. Now we’re going to ask you about **<u>what you ate</u>** in a **<u>regular week</u>**. We still want you to be thinking about **<u>the time since around late March.</u>**

During a regular week, how often did you eat meals from restaurants including delivery and takeout, instead of food prepared at home?

□ Never or rarely
□ 1-2 times per week
□ 3-4 times per week
□ 5-6 times per week
□ 1 time per day
□ 2 or more times per day

13. During a **<u>regular week since around late March</u>**, how often did you **<u>eat snacks (between meals)</u>** from a vending machine, snack bar, convenience store, gas station, or similar store outside of school property?

□ Never or rarely
□ 1-2 times per week
□ 3-4 times per week
□ 5-6 times per week
□ 1 time per day
□ 2 or more times per day

14. In a **<u>regular week since around late March</u>**, how often did you **<u>drink</u>** the following?

|  | Never<br>or<br>rarely | 1-2 times<br>per week | 3-4 times<br>per week | 5-6 times<br>per week | 1 time<br>per DAY | 2 times<br>per DAY | 3 or more<br>times<br>per DAY |
| --- | --- | --- | --- | --- | --- | --- | --- |
| **<u>Regular (sugar-sweetened) soda</u>** such as Coke, Pepsi, Sprite | ☐ | ☐ | ☐ | ☐ | ☐ | ☐ | ☐ |
| **<u>Diet (sugar-free) soda</u>** such as Diet Coke, Diet Pepsi, Diet Sprite | ☐ | ☐ | ☐ | ☐ | ☐ | ☐ | ☐ |
| **<u>Regular (sugar-sweetened) sports drinks</u>** such as Gatorade, Powerade | ☐ | ☐ | ☐ | ☐ | ☐ | ☐ | ☐ |
| **<u>Sugar-free sports drinks</u>** such as Gatorade Zero, Powerade Zero | ☐ | ☐ | ☐ | ☐ | ☐ | ☐ | ☐ |
| **<u>Other sugar-sweetened beverages</u>** such as sweetened tea, energy drinks, juice drinks like punch or lemonade | ☐ | ☐ | ☐ | ☐ | ☐ | ☐ | ☐ |
| **<u>Other sugar-free beverages</u>** such as sugar-free energy drinks, sugar-free lemonade | ☐ | ☐ | ☐ | ☐ | ☐ | ☐ | ☐ |
| **<u>100% Fruit juice</u>** such as 100% apple, orange, or grape juice, Juicy Juice | ☐ | ☐ | ☐ | ☐ | ☐ | ☐ | ☐ |
| **<u>Water</u>** (including tap water, bottled water or seltzer such as Dasani, Poland Spring, Aquafina) | ☐ | ☐ | ☐ | ☐ | ☐ | ☐ | ☐ |

15. In a regular week since around late March, how many days did you eat breakfast?

□ Never
□ 1 or 2 days each week
□ 3 or 4 days each week
□ 5 or 6 days each week
□ Every day

16. A serving of **<u>fruit or vegetables</u>** is about the size of your fist. During the past week…

|  | Less than 1<br>serving per day | 1 serving<br>per day | 2 servings<br>per day | 3 or 4<br>servings<br>per day | 5 or more<br>servings per<br>day |
| --- | --- | --- | --- | --- | --- |
| How many servings of **<u>fruit</u>** (fresh, frozen, or canned) did you eat on a usual day? | ☐ | ☐ | ☐ | ☐ | ☐ |
| How many servings of **<u>vegetables</u>** (fresh, frozen or canned) did you eat on a usual day? | ☐ | ☐ | ☐ | ☐ | ☐ |

17. Since around late March, have you gone on a diet to lose weight?

□ No
□ Yes

18. Now we’re going to ask you some questions about **<u>PHYSICAL ACTIVITY</u>**.

Physical Activity is any game, sport, or exercise that makes you move faster and breathe harder; makes your heart beat faster; and makes you sweat a little or a lot. We still want you to be thinking about the time **<u>since around late March</u>.**

During a **<u>regular week since around late March</u>**, how much time did you spend doing the following activities?

|  | None | Less than<br>½ hour a<br>week | ½-2<br>hours a<br>week | 2 ½-4<br>hours a<br>week | 4 ½-6<br>hours a<br>week | 6+ hours<br>a week |
| --- | --- | --- | --- | --- | --- | --- |
| **<u>VIGOROUS</u>** activity (Examples: jogging and running, biking hard, soccer, basketball, swimming laps) | ☐ | ☐ | ☐ | ☐ | ☐ | ☐ |
| **<u>MODERATE</u>** activity (Examples: walking quickly, easy bicycling, dancing, skateboarding, lifting weights, easy swimming, baseball, volleyball) | ☐ | ☐ | ☐ | ☐ | ☐ | ☐ |
| **<u>LIGHT</u>** activity (Examples: slow walking, shopping, bowling, golf, stretching) | ☐ | ☐ | ☐ | ☐ | ☐ | ☐ |

19. The next questions are about times you spent sitting/lying down while using your phone, tablet, computer, or gaming console. Again, we are talking about the time since around late March.

On a **regular WEEKDAY** (Monday-Friday), how many hours each day did you spend **sitting/lying down while**...

|  | None | Less than 1<br>hr a day | 1 hr a<br>day | 2 hrs<br>a day | 3 hrs<br>a day | 4 hrs<br>a day | 5+ hrs<br>a day |
| --- | --- | --- | --- | --- | --- | --- | --- |
| **Watching TV, DVD, or streaming videos**<br>(Hulu, Netflix, YouTube, Amazon, Vines,<br>cable). | ☐ | ☐ | ☐ | ☐ | ☐ | ☐ | ☐ |
| **Playing video games** (console, computer,<br>phone/tablet). | ☐ | ☐ | ☐ | ☐ | ☐ | ☐ | ☐ |
| **Browsing/posting/ messaging on social**<br>**media** (including Facebook, Instagram,<br>SnapChat, TikTok). | ☐ | ☐ | ☐ | ☐ | ☐ | ☐ | ☐ |
| **Communicating by voice or video chat**<br>(including Facetime, Skype, Facebook<br>Messenger, SnapChat, or others). | ☐ | ☐ | ☐ | ☐ | ☐ | ☐ | ☐ |
| **Browsing the web** (not for watching<br>videos, playing games, social media, or<br>homework). | ☐ | ☐ | ☐ | ☐ | ☐ | ☐ | ☐ |

20. On a **<u>usual SATURDAY OR SUNDAY</u>** (weekend) during a regular week, how many hours each day did you spend **<u>sitting/lying down while</u>**...

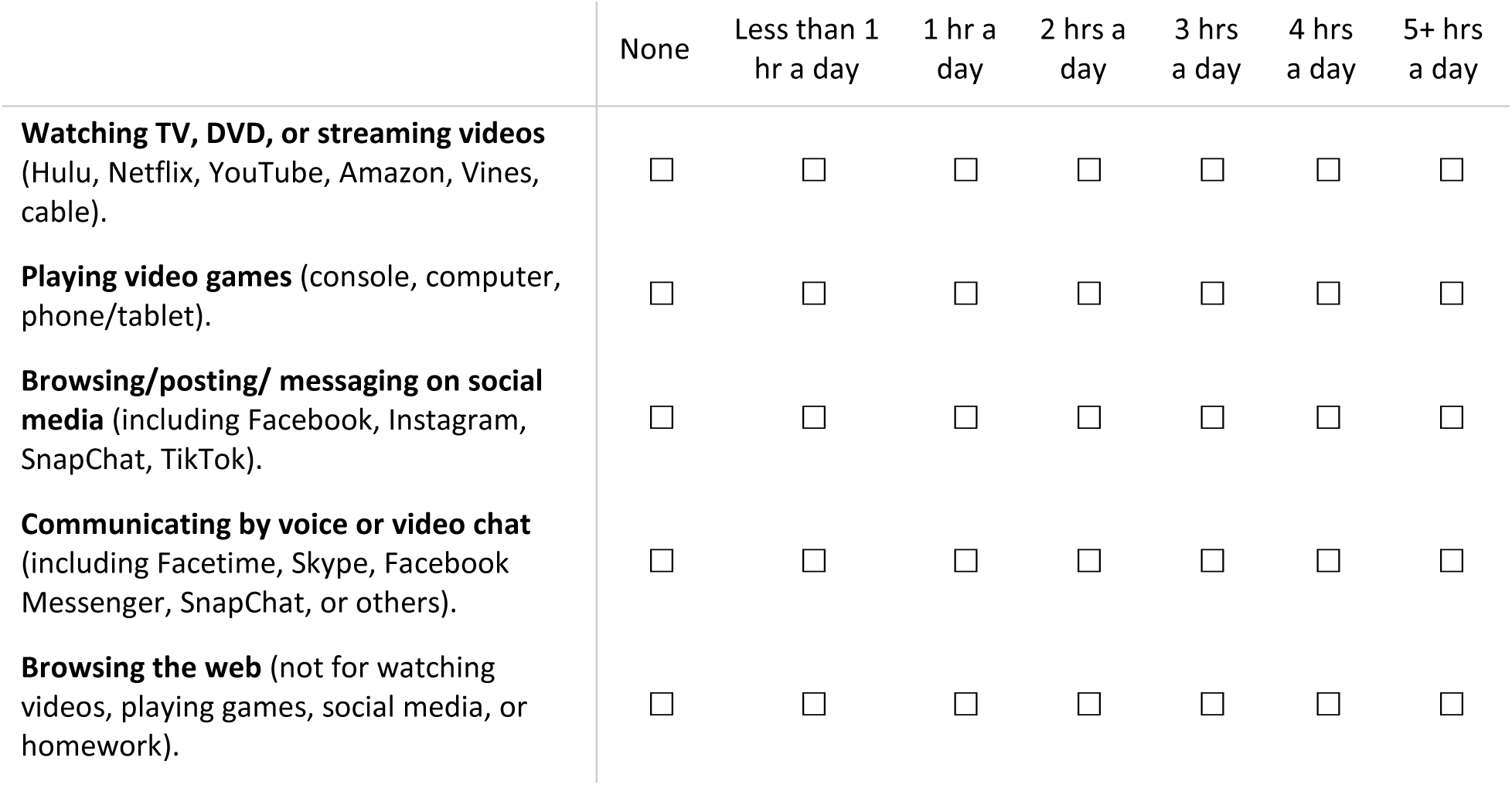

21. The next two questions are about times you spent playing **<u>active video games</u>** (where you’re standing up, dancing, jumping). Again, we are talking about the time **<u>since around late March</u>.**

On a **<u>regular WEEKDAY</u>** (Monday-Friday), how many hours per day did you spend doing these **<u>active/moving video games</u>?**

□ None
□ Less than 1 hour a day
□ 1 hour a day
□ 2 hours a day
□ 3 hours a day
□ 4 hours a day
□ 5+ hours a day

22. On a **<u>usual SATURDAY OR SUNDAY</u>** (weekend) during a regular week, how many hours each day did you spend **<u>doing these active/moving video games?</u>**

□ None
□ Less than 1 hour a day
□ 1 hour a day
□ 2 hours a day
□ 3 hours a day
□ 4 hours a day
□ 5+ hours a day

23. Check the boxes next to the 8th graders who are your **<u>friends</u>**.

<<ROSTER>>

**□ There is no one on this list I want to pick.**

24. If your friend is an 8th grader at this school, but is not included in the list on the previous page, write the name(s) below.

Please give first and last name:

______________________________

Please give first and last name:

______________________________

Please give first and last name:

______________________________

25. How many **<u>friends</u>**do you have who **<u>go to other schools</u>** who are AS CLOSE or CLOSER to you than the friends you picked in questions 23 and 24 above?

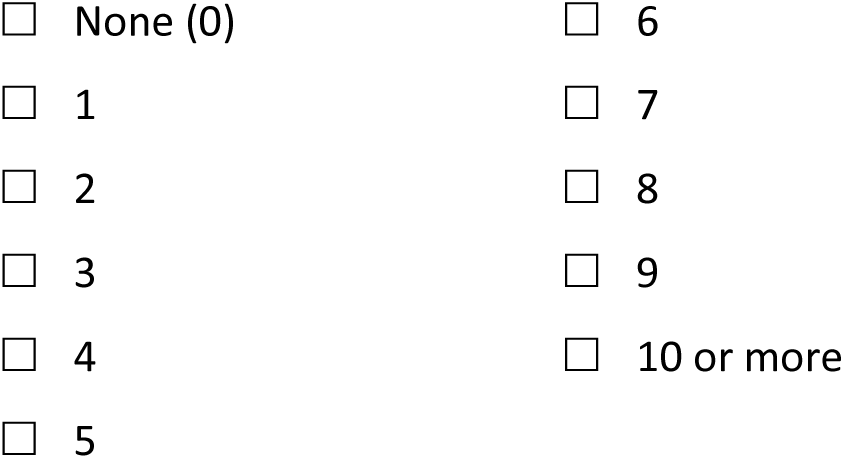

26. How many **<u>friends</u>** do you have in **<u>other grades at this school</u>** who are AS CLOSE or CLOSER to you than the friends you picked in questions 22 and 23 above?

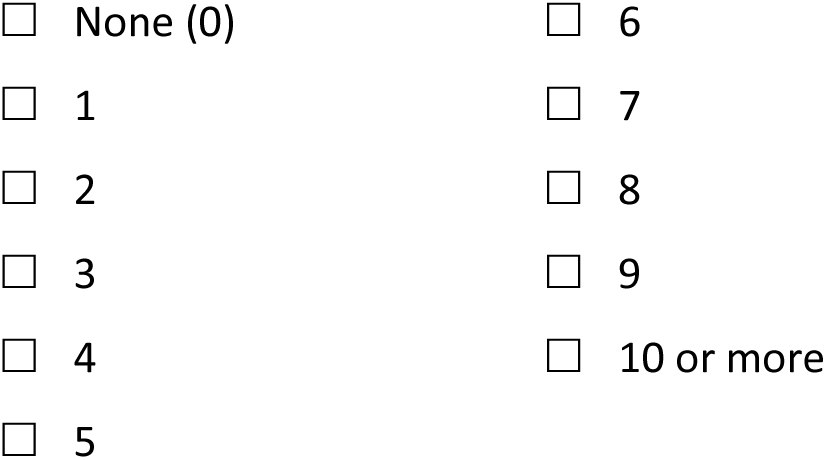

27. How strongly do you agree with the following statements? For these questions, think about your <u>friends</u>.

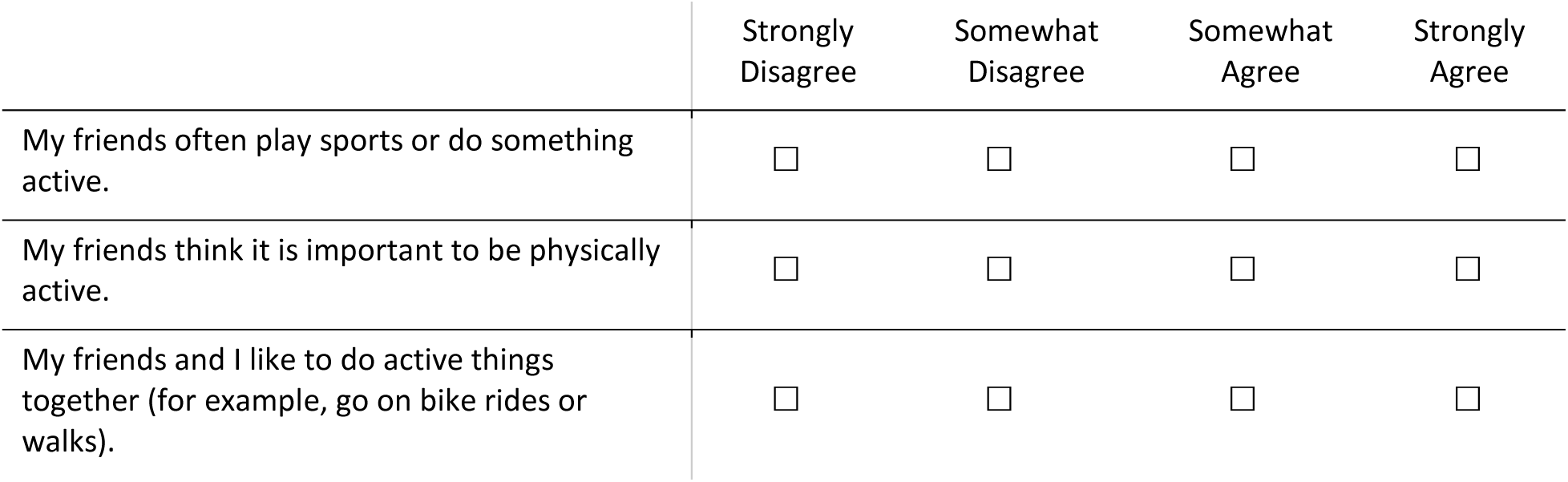

28. What is your gender?

□ Boy
□ Girl

Do you have anything else to add?

________________________________________

29. What is your birthdate?

Month _________

Day _________

Year _________

30. How do you describe yourself? Select ALL that apply. **(You can choose more than one.)**

□ American Indian or Alaska Native
□ Asian
□ Black or African American
□ Hispanic
□ Latino
□ Native Hawaiian or other Pacific Islander
□ White

31. Do you also describe yourself in any of the following ways? If so, select ALL that apply. **(You can choose more than one.)**

**If none of the choices fit, choose: *“I don’t describe myself in any of these ways.”***

|  |  |
| --- | --- |
| ☐ Puerto Rican | ☐ Cuban |
| ☐ Dominican | ☐ Guatemalan |
| ☐ Brazilian | ☐ Jamaican |
| ☐ Salvadorian | ☐ Mexican |
| ☐ Venezuelan | ☐ Panamanian |
| ☐ Bahamian | ☐ Peruvian |
| ☐ Other _____ |  |
| ☐ I don't describe myself in any of these ways. |  |

32. Do you consider yourself multiracial or mixed race?

□ No
□ Yes

33. Do you have anything more to add about how you describe yourself? ___________________________________________________________ ___________________________________________________________ ___________________________________________________________ ___________________________________________________________

34. How many working computers (desktop, laptop, or tablet) does your family own? Do not count phones.

□ None
□ One
□ Two
□ Three
□ Four or more

35. Does your family own a vehicle (car, van, or truck)?

□ None
□ One
□ Two or more

36. During the past 12 months, how many times did you travel for vacation (on an airplane or train) with your family?

□ Not at all.
□ Once.
□ Twice.
□ More than twice.

37. Do you have your own bedroom for yourself?

□ No.
□ Yes.

38. Does your family have their own (clothes) washing machine in your home that is not shared with another household?

□ No.
□ Yes.

39. How many bathrooms are in your home?

□ One.
□ Two.
□ More than two.

40. For these questions, think about **<u>the home where you live most of the time</u>**. Does your mother/step mother in your home **<u>work at a job</u>**?

□ Yes
□ No
□ I don’t know
□ There is no mother/step mother in my home

40a. If you answered “yes” to Question 40, could you tell us what your mother’s/step mother’s job is? If you don’t know the name of the job, describe the type of work as closely as you can.

_______________________________________________________

_______________________________________________________

40b. **<u>How far in school</u>** did your mother/step mother go?

□ Did not graduate from high school
□ Graduated from high school
□ Started college but did not graduate
□ Graduated from college

41. For these questions, think about **<u>the home where you live most of the time</u>**. Does your father/step father in your home **<u>work at a job</u>**?

□ Yes
□ No
□ I don’t know
□ There is no father/step father in my home

41a. If you answered “yes” to Question 41, could you tell us what your father’s/step father’s job is? If you don’t know the name of the job, describe the type of work as closely as you can. _______________________________________________________ _______________________________________________________

41b. **<u>How far in school</u>** did your father/step father go?
  □ Did not graduate from high school
  □ Graduated from high school
  □ Started college but did not graduate
  □ Graduated from college

42. Is there anything else you’d like to share about either of your parents’ jobs or education?

_______________________________________________________

_______________________________________________________

43. Now we’re going to ask you a few questions about your family and your health. For these questions, think about the home where you live most of the time.

Which of the following people also live in the same household with you? (Mark all that apply.)

□ Father (or male guardian).
□ Mother (or female guardian).
□ Siblings (brothers or sisters).
□ Grandparents.
□ Other relative(s).
□ Non-relative(s).

44. How many brothers and sisters do you have? (Include stepbrothers and stepsisters and half-brothers and half-sisters)

|  | None | 1 | 2 | 3 | 4 | 5 | 6+ |
| --- | --- | --- | --- | --- | --- | --- | --- |
| Older brothers and sisters | ☐ | ☐ | ☐ | ☐ | ☐ | ☐ | ☐ |
| Younger brothers and sisters | ☐ | ☐ | ☐ | ☐ | ☐ | ☐ | ☐ |

45. Is there anyone in the 8th grade that is your **<u>cousin, sibling, or lives with you</u>**?

If so, please give their first and last name, and select/circle any options below that show who they are to you.

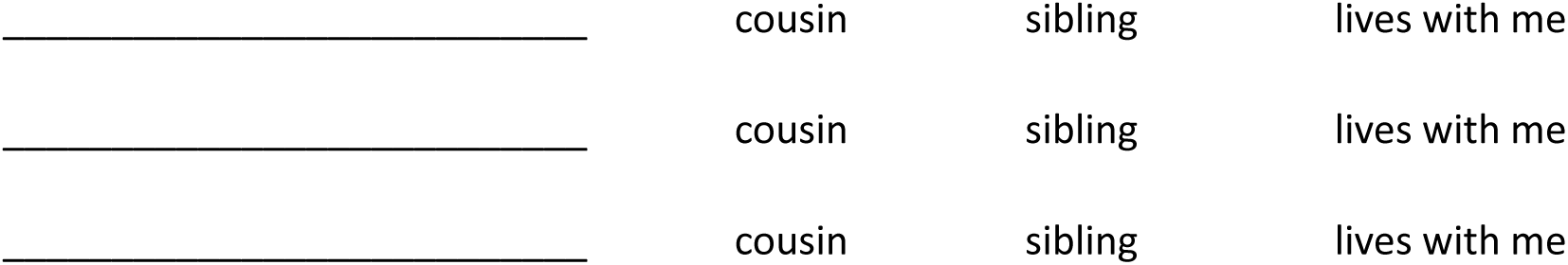
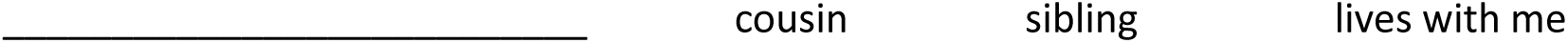

|  |  |  |  |
| --- | --- | --- | --- |
| _____ | cousin | sibling | lives with me |
| _____ | cousin | sibling | lives with me |
| _____ | cousin | sibling | lives with me |

47. How **<u>tall</u>** are you, without shoes?

Your height:______feet______inches

48. How much do you **<u>weigh (in pounds)</u>**?

Your weight:_______pounds

49. **<u>During a regular week since around late March</u>**, how much time did you spend with other 8th graders from your school **<u>outside of school hours</u>**?

|  | None | Less than 1/2 hour a week | ½-2 hours a week | 2 ½-4 hours a week | 4 ½-6 hours a week | 6+ hours a week |
| --- | --- | --- | --- | --- | --- | --- |
| In person | ☐ | ☐ | ☐ | ☐ | ☐ | ☐ |
| Online | ☐ | ☐ | ☐ | ☐ | ☐ | ☐ |

50. <u>During a regular week since</u> <u>around late March</u>, how much time did you spend with students who go to <u>other schools</u> (or who are in <u>other grades</u> at your school) <u>outside of school hours</u>?

|  | None | Less than 1/2 hour a week | ½-2 hours a week | 2 ½-4 hours a week | 4 ½-6 hours a week | 6+ hours a week |
| --- | --- | --- | --- | --- | --- | --- |
| In person | ☐ | ☐ | ☐ | ☐ | ☐ | ☐ |
| Online | ☐ | ☐ | ☐ | ☐ | ☐ | ☐ |

51. Please use the space below to tell us anything else about **<u>how your social life has changed,</u> <u>since around late March.</u>**

___________________________________________________________

52. During a regular week **<u>since around late March</u>**, what time did you usually **go to bed** in the evening (turn out the lights in order to go to sleep)?

|  | **Weekday**<br>**(Check one box)** | **Weekend**<br>**(Check one box)** |
| --- | --- | --- |
| 8:00 pm or earlier | ☐ | ☐ |
| 8:30 pm | ☐ | ☐ |
| 9:00 pm | ☐ | ☐ |
| 9:30 pm | ☐ | ☐ |
| 10:00 pm | ☐ | ☐ |
| 10:30 pm | ☐ | ☐ |
| 11:00 pm | ☐ | ☐ |
| 11:30 pm | ☐ | ☐ |
| 12:00 am (midnight) | ☐ | ☐ |
| 12:30 am | ☐ | ☐ |
| 1:00 am | ☐ | ☐ |
| 1:30 am | ☐ | ☐ |
| 2:00 am or later | ☐ | ☐ |

53. During a regular week **<u>since around late March</u>**, what time did you usually **get out of bed** in the morning?

52. During a regular week since around late March, what time did you usually **get out of bed** in the morning?
|  | Weekday<br>(check one box) | Weekend<br>(check one box) |
| --- | --- | --- |
| 5:00 am or earlier | ☐ | ☐ |
| 5:30 am | ☐ | ☐ |
| 6:00 am | ☐ | ☐ |
| 6:30 am | ☐ | ☐ |
| 7:00 am | ☐ | ☐ |
| 7:30 am | ☐ | ☐ |
| 8:00 am | ☐ | ☐ |
| 8:30 am | ☐ | ☐ |
| 9:00 am | ☐ | ☐ |
| 9:30 am | ☐ | ☐ |
| 10:00 am | ☐ | ☐ |
| 10:30 am | ☐ | ☐ |
| 11:00 am | ☐ | ☐ |
| 11:30 am | ☐ | ☐ |
| 12:00 pm (noon) or later | ☐ | ☐ |

Being directly exposed to COVID-19 may also affect your eating, physical activity, and screen time. Remember, we keep all of your answers private, and you can skip any question you don’t feel comfortable answering.

54. **<u>Since around late March</u>**, were you **<u>exposed to COVID-19</u>** by being in close proximity to a person diagnosed with (or tested positive for) COVID-19?

□ Not that I know of
□ I think so
□ Yes
Since around late March**, were *you*** diagnosed with (or tested positive for) COVID-19**?**

□ No
□ Yes
If you could get a COVID-19 vaccine now, would you want it?

□ Definitely not
□ Probably not
□ Might or might not
□ Probably yes
□ Definitely yes

55. Anything else you’d like to add about the COVID-19 vaccine or your exposure to COVID-19?
____________________________________________________________________

56. Imagine that you belong to a **<u>friendship group</u>** (**a group of three or more friends**). For you, how important is it that others in the friendship group **<u>agree with you</u>** on who is **part of** the group, or **out of** the group?
  □ Not at all important
  □ Slightly important
  □ Moderately important
  □ Very important
  □ Extremely important

57. Imagine that this ladder is a way of picturing your school.
  - At the top of the ladder are the students in your school who most others like, respect, and want to hang around with.
  - At the bottom are students who no one likes, respects, or wants to hang around with.

Where would you place yourself on this ladder?

**Tap/Put an X** on the ONE step that best represents where you would be on this ladder.

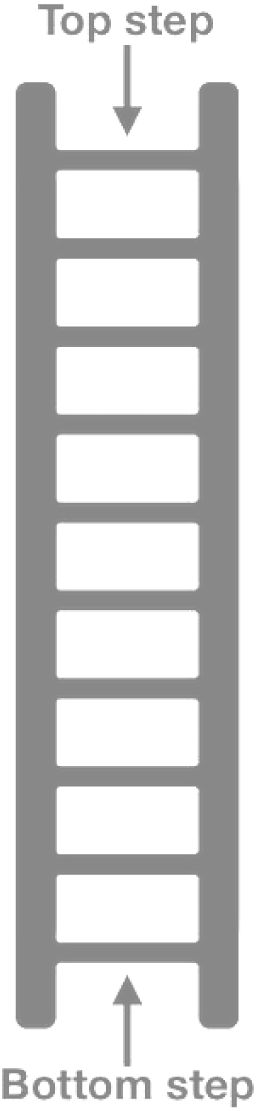

58. What languages do you speak? (Choose all that apply)

□ English
□ Spanish
□ Other:________

59. Which language do you speak most often **<u>at home</u>**? (Choose only one)
  □ English
  □ Spanish
  □ Other:________

60. Do you have any final comments about how you answered this survey? Please let us know below. ________________________________________________________ ________________________________________________________

61. What school do you go to? ____________________________________________

*Thank you so much for your time and for continuing to take part in our study. We really appreciate your time and help*.

62. Please write in your street address below so we can mail your gift card to the right place!

________________________________________________________

________________________________________________________

________________________________________________________

63. Almost done! To be sure our records are accurate, check the box next to YOUR name one last time from the list below.

<< ROSTER >>

**□ I can’t find my name on the list above!**

If you’re sure your name isn’t on the list above, write it in here (first & last).

______________________________________________

